# Multi-ancestry omic Mendelian randomization revealing putative drug targets of COVID-19 severity

**DOI:** 10.1101/2020.05.07.20093286

**Authors:** Jie Zheng, Yuemiao Zhang, Huiling Zhao, Yi Liu, Denis Baird, Mohd Anisul Karim, Maya Ghoussaini, Jeremy Schwartzentruber, Ian Dunham, Benjamin Elsworth, Katherine Roberts, Hannah Compton, Felix Miller-Molloy, Xingzi Liu, Lin Wang, Hong Zhang, George Davey Smith, Tom R Gaunt

## Abstract

Recent omic studies prioritised several drug targets associated with coronavirus disease 2019 (COVID-19) severity. However, little evidence was provided to systematically estimate the effect of drug targets on COVID-19 severity in multiple ancestries. In this study, we applied Mendelian randomization (MR) and colocalization approaches to understand the putative causal effects of 16,059 transcripts and 1,608 proteins on COVID-19 severity in European and effects of 610 proteins on COVID-19 severity in African ancestry. We further integrated genetics, clinical and literature evidence to prioritised additional drug targets. Additional sensitivity analyses including multi-trait colocalization and phenome-wide MR were conducted to test for MR assumptions.

MR and colocalization prioritized four protein targets, FCRL3, ICAM5, ENTPD5 and OAS1 that showed effect on COVID-19 severity only in European ancestry and one protein target, SERPINA1, only showed effect in African ancestry (odds ratio [OR] in Africans=0.369, 95%CI=0.203 to 0.668, P=9.96×10^−4^; OR in Europeans=1.021, P=0.745). One protein, ICAM1, showed suggestive effect on COVID-19 severity in both ancestries (OR in Europeans=1.152, 95%CI=1.063 to 1.249, P=5.94×10^−4^; OR in Africans=1.481, 95%CI=1.008 to 2.176; P=0.045). The phenome-wide MR of the prioritised targets on 622 complex traits identified 726 potential causal effects on other diseases, providing information on potential beneficial and adverse effects. Our study prioritised six proteins as potential drug targets for COVID-19 severity. Several of them were targets of existing drug under trials of COVID-19 or related to the immune system. Most of these targets showed different effects in European and African ancestries, which highlights the value of multi-ancestry MR in informing the generalizability of COVID-19 drug targets across ancestries. This study provides a first step towards clinical investigation on COVID-19 and other types of coronaviruses.

**Research in context:** *Evidence before this study:* We searched key terms in PUBMED published before Feb 1st 2022, with the terms: (“COVID-19, “coronavirus”) AND (“omics” or “protein” or “transcript”) AND (“Genome-wide association study” or “Mendelian randomization”). We found multiple studies identified targeted genes or proteins associated with COVID-19. However, there is little human genetics evidence support the ancestry-consistent or ancestry-specific genes/proteins associated with COVID-19.

*Added value of this study:* To our knowledge, this is the first comprehensive genetic study that identified protein targets that showed effect on COVID-19 severity in European and African ancestries. Our study identified one protein, SERPINA1, that showed effects on COVID-19 in African ancestry (OR=0.369, P=9.96×10^−4^), but not in European ancestry (OR=1.021, P=0.745). In addition, our study identified four additional protein targets, FCRL3, ICAM5, ENTPD5 and OAS1, that showed effect on COVID-19 severity in Europeans. One protein ICAM1 showed suggestive effect in both ancestries. Some of these proteins are related to the immune system and/or are targets of existing drug under trials of COVID-19.

*Implications of all available evidence:* Our study prioritised six drug targets for COVID-19 severity, five of them showed different effects in European and African ancestries. This suggested that drug targets may have different responses on COVID-19 severity in different ancestries. Our study also highlights the value of intercellular adhesion molecule (ICAM) family in relation with COVID-19 severity in both ancestries.

## Introduction

The outbreak of Severe Acute Respiratory Syndrome Coronavirus 2 (SARS-CoV-2), the causative agent of novel coronavirus disease 2019 (COVID-19), has been a global pandemic. It has infected a large portion of the world population with a wide spectrum of manifestations, ranging from completely asymptomatic carriers to critical respiratory failure, multi-organ dysfunction, and death^1^. This heterogeneity may relate to different host responses involving multiple human proteins and pathways. Considering the relatively high mortality rate of patients with severe disease^2^, search for optimal treatments, such as novel drugs to improve survival rate of severe patients is a key objective to reduce the impact of both the current and potential future coronavirus epidemics.

Currently, some clinical trials have showed beneficial effect of some approved drugs on COVID-19, including IL6R antagonist^3^ and ACE inhibitor^4^. Some experimental studies identified a set of human proteins interacted with SARS-CoV-2^5^, which may provide drug targets for general coronavirus interventions. Some genetic and epidemiology studies suggested that drug response of COVID-19 was different among ethnic groups. For example, Ammar et al showed that IL6R inhibition are likely to showed 3.4 times more impaired response in sub-Saharan Africans compared to East Asians and 1.8 times compared to Europeans^6^. To improve the generalizability of drug interventions for COVID-19 severity, we need to use some new approaches to predict treatment response in different ancestries.

Omics data could play a role in supporting efficient drug development for various diseases including COVID-19. Recent studies have begun to support the role of genetics in predicting drug trial success^7^. Some multi-omics studies have further demonstrated the value of molecular quantitative trait locus (QTL) studies in repurposing existing targets to additional indications as well as prioritizing novel drug targets^8^. One approach to utilising molecular QTL data is Mendelian randomization (MR)^9^. MR uses genetic variants as instrumental variables to estimate the effect of an exposure (e.g. measured levels of a protein) on an outcome (e.g. COVID-19 severity), which could prioritise drug targets cost-effectively. Recent multi-ancestry genetic studies have further showcased the value of omics analyses in predicting treatment response in various populations^10,11^.

Recent GWAS^12,13,14^ and MR studies^15,16,17^ utilising data from the COVID-19 Host Genetics Initiative (HGI; https://www.covid19hg.org/) and GenOMICC consortium have identified a set of genes (e.g. *ABO, OAS1, IFNAR2, IL10RB*) associated with various COVID-19 phenotypes. However, there are issues that require consideration: i) some genes and proteins (e.g. ABO) are well-known to be pleiotropic, which may violate the “no pleiotropy” (or “exclusion restriction”) assumption of MR; ii) when comparing severe COVID-19 cases with population controls as an outcome, it is not possible to separate the causal effects of becoming infected from any causal effects on disease progression after infection (despite these potentially being separate mechanisms)^18^; iii) collider bias (also known as selection bias, sampling bias or ascertainment bias)^19^. Each of these could induce spurious associations between the target and COVID-19. Careful instrument and outcome selection are needed for the drug target MR of COVID-19.

The aim of this study is to prioritise potential drug targets for COVID-19 severity as well as to identify potential beneficial and adverse effects of these targets on other diseases. We combined genetic association information of 16,059 transcripts and 2,324 proteins in European ancestry^8,20,21^ and 610 proteins in African ancestry^10^, and applied a recent omics MR analysis pipeline^11^ to estimate the causal effects of these targets on COVID-19 severity in the two ancestries separately. To enable rapid queries, results of all analyses are available in an open access online platform (https://epigraphdb.org/covid-19/ctda/).

## Methods

### Study design and participants

**Figure 1** described the study design of this study. We aimed to prioritise drug targets for COVID-19 severity in European and African ancestries. Three sets of exposures that proxying drug target effects have been setup in this study: i) expression levels of 1,608 proteins (N≤7,212; **Table S1A**) and 16,059 genes (N≤31,684; **Table S2**) from European ancestry; ii) expression level of 610 proteins from African ancestry (N≤1,871; **Table S1B**); iii) tissue-specific gene expression of 353 genes with literature evidence of associated with COVID-19 (**Table S3-5**). The outcome was COVID-19 severity from two studies (HGI N cases=928, N controls=2028; GenOMICC Europeans N cases=1,676, N controls=1,676; GenOMICC Africans N cases=190, N controls=190; **Table S6**). The putative causal effects of the selected drug targets on COVID-19 severity were estimated using MR and colocalization in European and African ancestries separately. Several sensitivity analyses including multi-trait colocalization and phenome-wide MR were conducted to validate core MR assumptions.

**Figure 1.**
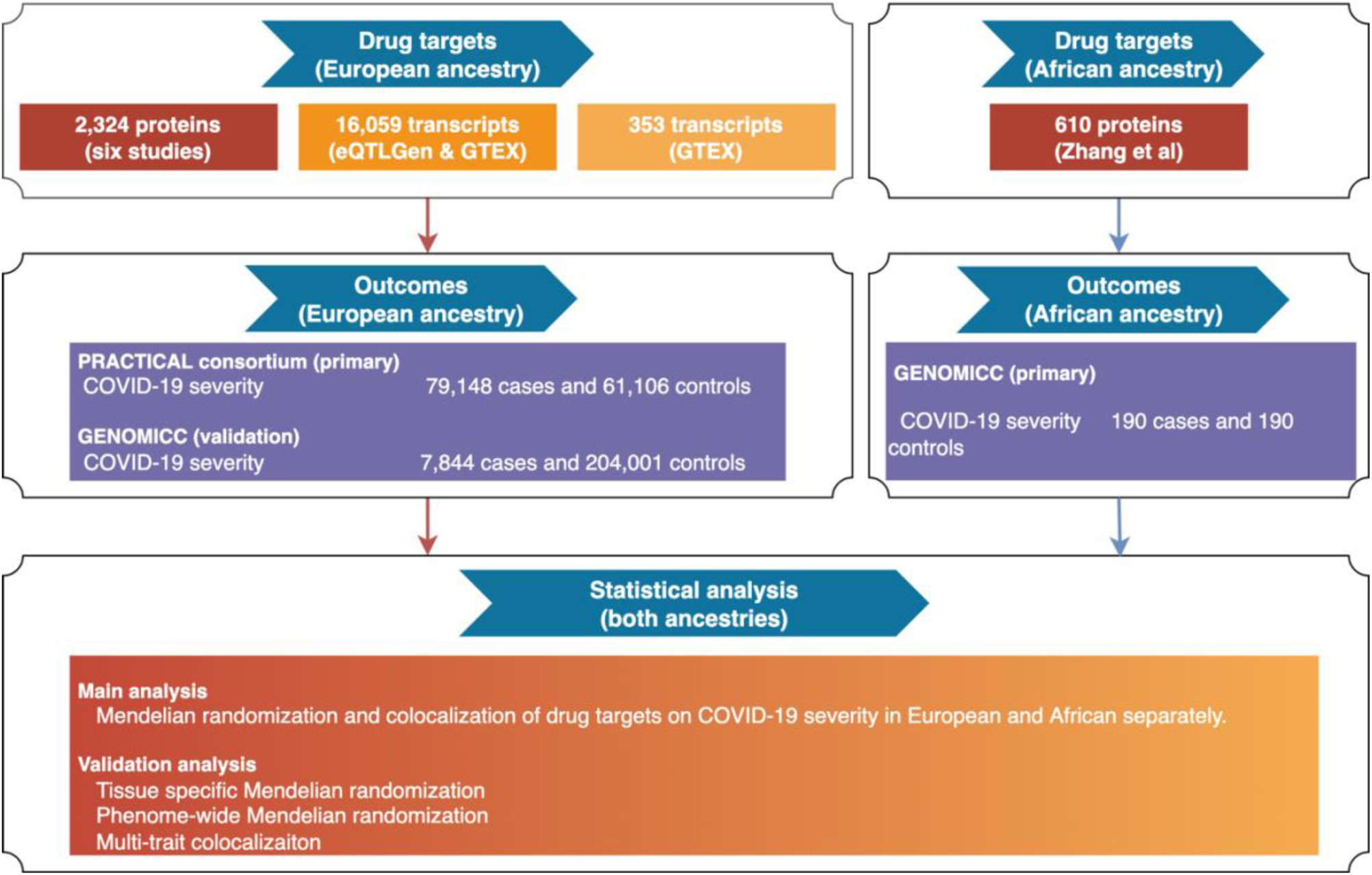
Study design of the multi-ancestry Mendelian randomization study to prioritise drug targets for COVID-19 severity.

### Instruments selection for drug targets

Proteins and transcripts of genes were selected as potential drug targets in this study since they are direct targets for most of the approved drugs. To generate genetic instruments for potential drug targets, genetic variants robustly associated with the expression of proteins and genes (with P<5×10^−8^), irrespective of genomic position of variants, were selected from six proteome datasets implemented in two studies^8,10^ and two transcriptome datasets^20,21^.

As showed in **Figure 1**, three sets of genetic instruments that proxying drug target effects were selected in this study: i) 7,092 instruments of 1,608 proteins and 39,630 instruments of 16,059 gene expressions in the European ancestry; ii) 3,550 instruments of 610 proteins in the African ancestry; iii) 1,493 instruments of 353 COVID-19 related drug targets from literatures (**Table S4**).

### Outcome selection

The genetic associations of COVID-19 severity were obtained from the COVID-19 HGI (https://www.covid19hg.org/). Among the six available genetic association data listed in **Table S6**, we considered the hospitalized COVID-19 vs. not hospitalized COVID-19 as the most suitable dataset to represent COVID-19 severity. The major advantage of this data compared to other datasets (e.g. COVID-19 vs. population) is that all participants of this genetic analysis were confirmed COVID-19 cases, therefore this analysis will be less bias by data selection (e.g. there will be some unconfirmed COVID-19 cases in the population so it is not a perfect control dataset). We used the GWAS summary statistics of the hospitalized COVID-19 vs. not hospitalized COVID-19 as the outcome for the MR analysis. This GWAS combined European samples from three large scale biobanks: UK Biobank, DECODE and FinnGen (928 confirmed hospitalized COVID-19 patients, 2,028 confirmed non hospitalised COVID-19 patients). To validate and identify additional causal genes and/or proteins, we used the COVID-19 severity dataset from the GENOMICC study^13^. Both European- and African-specific datasets of COVID-19 severity were used in this study (**Table S6**).

### Statistical analysis

#### Mendelian randomization analysis

In the main analysis, the effects of selected proteins and transcripts were tested against COVID-19 severity using MR in European (HGI data as discovery and GENOMICC as validation) and African ancestries (GENOMICC data as discovery) separately. The Wald ratio method was used to obtain MR effects for protein and/or transcript targets with only one instrument. For protein and/or transcript targets with two or more instruments, the inverse variance weighted (IVW) method was used to estimate the MR effects.

#### Colocalization analysis

Genetically predicted associations between a protein or gene and COVID-19 severity may arise any of the four scenarios: causality, reverse causality, confounding by linkage disequilibrium (LD) between the leading SNPs for proteins and phenotypes or horizontal pleiotropy (**Figure S1**). For drug targets showed robust MR effects on COVID-19 severity in the main MR analysis, (Bonferroni corrected threshold, P for protein< 5.97×10^−5^ or P for transcript< 3.56×10^−6^), we conducted single-trait colocalization^22^ to distinguish causality from confounding by LD (using the “coloc” R package). This approach estimates the posterior probability of each genomic locus containing a single variant affecting both the target and the phenotype^22^. We used the default prior probabilities that a variant is equally associated with each phenotype (p1=1×10^−4^; p2=1×10^−4^) and both phenotypes jointly (p12=1×10^−5^). A posterior probability of > 70% for the colocalization hypothesis in this analysis would suggest that the two association signals are likely to colocalize within the test region (noted as “Colocalised”). The rest of the target-phenotype associations were noted as “Not colocalised”.

#### Tissue Specificity analysis

The functional receptor of SARS-CoV-2, ACE2, is highly expressed in multiple organs, including gastrointestinal tract, gallbladder, testis, and kidney. This is consistent with the fact that whilst SARS-CoV-2 infection primarily manifests with acute respiratory illness. The presence of SARS-CoV-2 in the alimentary tract for longer than in the respiratory system^23^ suggests that the intestine may be a hidden reservoir of SARS-CoV-2. We therefore set out to explore differences in potential target effects in different tissues. In order to understand the tissue specific effects of the candidate targets of COVID-19 on human phenotypes, we selected the 9 tissues in which ACE2 is highly expressed, including testis, lung, kidney cortex, kidney glomerular, kidney tubulointerstitial, stomach, colon transverse, small intestine terminal ileum and colon sigmoid.

Tissue specific gene expression data of the 353 targets in each selected tissue were obtained from GTEx V8^21^. After selection, 580 instruments of 218 gene transcripts were selected in the 9 tissues, which included 141 instruments for 132 gene transcripts in testis, 125 instruments for 115 transcripts in lung, 20 instruments for 20 transcripts in kidney cortex, 6 instruments for 6 in kidney glomerular, 8 instruments for 8 transcripts in kidney tubulointerstitial, 71 instruments for 67 transcripts in stomach, 84 instruments for 81 transcripts in colon sigmoid, 98 instruments for 96 transcripts in colon transverse and 41 instruments for 39 transcripts in small intestine terminal ileum (**Table S5**). The same MR and colocalization analysis pipeline were applied for the tissue specific analysis.

#### Multi-trait colocalization analysis of prioritised targets

For protein and/or transcript targets with robust MR (P_protein< 5.97×10^−5^ or P_transcript< 3.56×10^−6^) and colocalization (probability >80%) evidence, we explored whether the causal variants were shared across transcriptome, proteome and COVID-19 severity for top MR findings. We applied multi-trait colocalization implemented in the moloc R package^24^. The default prior probabilities of 1×10^−4^ for any one layer of association, 1×10^−6^ for any two layers of associations and 1×10^−7^ for colocalization of all three layers of associations were used in the moloc analysis. An overall colocalization probability of three traits (Pa,bc+Pab,c+Pac,b+Pabc) > 80% would suggest that the three association signals are likely to colocalize within the test region.

#### MR PheWAS of prioritised targets for COVID-19 severity

For the 353 prioritised drug targets with trial or experimental evidence and targets with robust MR/colocalization evidence of association with COVID-19 severity, we further conducted a phenome-wide MR analysis (MR-PheWAS) to identify potential beneficial and/or adverse effects of these targets on other human diseases. The QTLs for the prioritised targets were chosen as the exposures for the MR-PheWAS. For the outcomes, 49 viral infection phenotypes from the GWAS Catalog (https://www.ebi.ac.uk/gwas/downloads/summary-statistics), 504 human diseases and 72 disease related traits (e.g. blood lipids) selected from the IEU Open GWAS database^25^ were selected as outcomes for this analysis (**Table S7**). These 573 outcomes were selected using the following inclusion criteria:

- The GWAS with the greatest expected statistical power (e.g. largest sample size / number of cases) when multiple GWAS records of the same disease / risk factor were available in the Open GWAS database.
- GWAS with betas, standard errors and effect alleles for all tested variants (i.e. full GWAS summary statistics available)

#### Analysis software

The MR analyses (including Wald ratio, IVW, two-step MR and bidirectional MR) were conducted using the TwoSampleMR R package (github.com/MRCIEU/TwoSampleMR)^26^. The MR results were plotted as Manhattan plots and forest plots using code derived from the ggplot2 package in R (https://cran.r-project.org/web/packages/ggplot2/index.html).

## Results

### Estimated causal effects of targets on COVID-19 severity

For the main MR analysis in European ancestry, the protein level of ENTPD5 showed a positive effect on COVID-19 severity after multiple testing correction (odds ratio [OR] of COVID-19 severity per standard deviation change of protein level= 2.07, 95%CI=1.47 to 2.92, P=3.29×10^−5^; **Table S8A**) using data from HGI. The protein level of OAS1 (OR=0.440, 95%CI=0.315 to 0.615, P= 1.57×10^−6^), FCRL3 (OR=1.032, 95%CI=1.030 to 1.034, P=2.40×10^−191^) and ICAM5 (OR=0.780, 95%CI=0.691 to 0.880, P=5.74×10^−6^) showed MR association with COVID-19 severity (**Table S8B**) using data from GENOMICC. As a positive control, inhibition of IL6R showed a protective effect on COVID-19 severity in European ancestry (OR=0.890, P=2.84×10^−^ 3), but the effect was much weaker in African ancestry (OR=1.351, 95%CI=0.892 to 2.045, P=0.155). For the MR analysis on transcripts, none of the transcript showed robust MR evidence using data from HGI (**Table S9A**), while expression of two genes, *LZTFL1* and *SLC4A10*, showed robust MR evidence after correction for multiple testing (**Table S9B**).

For the main MR analysis in African ancestry, the protein level of SERPINA1 showed a robust effect on COVID-19 severity in African ancestry (OR=0.369, 95%CI=0.203 to 0.668, P=9.96×10^−^ 4), but very unlikely to show an effect in European ancestry (OR=1.021, 95%CI=0.901 to 1.157, P=0.745).Using a more relaxing P-value threshold of 0.05, we found that expression level of one additional protein, ICAM1, showed robust effect on COVID-19 severity in European (OR=1.152, 95%CI=1.063 to 1.249, P=5.94×10^−4^; **Table S8B**) and suggestive effect in African (OR=1.481, 95%CI=1.008 to 2.176, P=0.045; **Table S8C**) ancestries.

For the 353 prioritised targets with experimental or trial evidence, all of them showed no strong MR evidence of a causal effect on COVID-19 severity. Only expression of 35 genes and levels of one protein (NPC2) showed nominal association on COVID-19 severity (we used a lenient P value threshold of P < 0.05, maximizing the number of possible genes analysed but also allowing readers to filter out associations should they wish to apply a more stringent threshold, **Table S10A** and **S10B**). All of these 36 targets were reported to interact with SARS-CoV-2 proteins in human cell lines (**Table S4**).

For the six targets, with strong MR evidence of a causal effect on COVID-19 severity, we conducted single-trait colocalization to distinguish causality from confounding by LD. This analysis showed evidence of a shared genetic effect between protein levels of ENTPD5 (colocalization probability [*p*]=99.9%), OAS1 (*p*=95.8%) FCRL3 (*p*=100%), ICAM5 (*p*=78.1%) and on COVID-19 severity in European ancestry and potential shared effect of protein level of SERPINA1 on COVID-19 severity in African ancestry (*p*=64.3%) (**Table S11A)**.

### Shared causal effect among gene expressions and protein levels of prioritised drug targets and COVID-19 severity

We observed strong MR (P=5.66×10^−5^) and colocalization (probability=99.9%) evidence to support the effect of gene expression level of *ENTPD5* on COVID-19 severity (**Table S11B**). The multi-trait colocalization analysis suggested robust colocalization evidence to support shared genetic aetiology of the three traits (shared probability=95.4%; **Figure 2**; **Table S11C**), which further strengthens the evidence level of the association between ENTPD5 and COVID-19 severity. For the OAS1, we observed little evidence of causal effect of the gene expression level of OAS1 on COVID-19 severity (P=0.568) (**Table S11B**).

**Figure 2.**
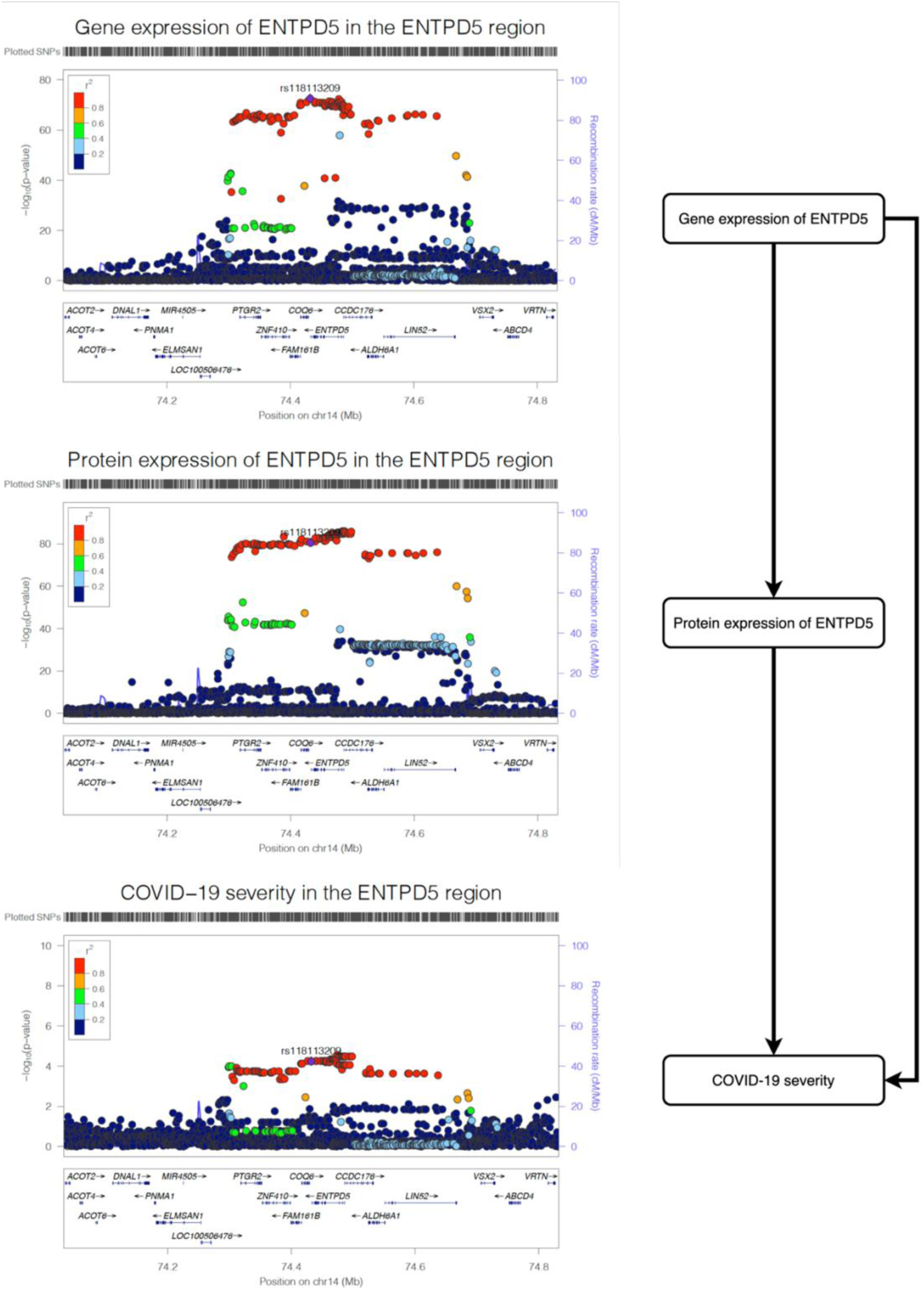
Regional plot of gene expression of *ENTPD5*, protein expression of ENTPD5 and COVID-19 severity in the *ENTPD5* region. The three regional plots refer to the genetic associations of (A), gene expression of *ENTPD5*; (B) protein expression of ENTPD5; (C) and COVID-19 severity. The X-axis is the chromosome and position of the *ENTPD5* region.

### Estimated beneficial and adverse effects of the prioritised targets on other complex diseases

We estimated potential beneficial and/or adverse effects of the protein and gene expression level of the prioritised targets on 622 traits (**Table S7**) using MR-PheWAS. 45,938 target-disease associations were tested in plasma proteome and/or transcriptome in whole blood (P<1.09×10^−6^ at a Bonferroni-corrected threshold). Where data was available, we also tested the tissue-specific effects of gene expression of the same targets on the outcome phenotypes using data from GTEX. Overall, 372,830 target-disease associations were estimated in the 11 COVID-19 relevant tissues.

As showed in **Figure S2**, protein level of ENTPD5 was not strongly associated with any of the 622 traits. Considering a more inclusive threshold (a lenient P value threshold of P<0.05), the MR-PheWAS suggested that reducing protein levels of ENTPD5 may increase the risk of lipid metabolism disorders (OR=0.93, 95%CI=0.90 to 0.97, P=1.41×10^−4^) and may increase the risk of joint disorders such as arthropathies (OR= 0.94, 95%CI=0.91 to 0.98, P= 0.002; **Figure S2** and **Table S12A**). The MR-PheWAS also suggested that genetically predicted ENTPD5 expression level was not associated with any disease (e.g. asthma) that might influence likelihood of seeking COVID-19 testing (the collider) (see **Figure S3** for more information).

For the 353 targets prioritised by literature evidence, we observed 833 target-disease associations with robust MR evidence in the 11 tested tissues. Using the same omics data as the MR analysis, 726 of the 833 (87.2%) associations also showed strong colocalization evidence (colocalization probability > 80%) (**Table S12B**), making these as more reliable findings of this study.

## Discussion

Genetic study of omics, provide a cost-effective approach to prioritise drug targets and predict drug response in multiple ancestries^8,11^. In this study, we applied omics MR in multiple ancestries to prioritise drug targets for COVID-19 severity. Our omics MR and colocalization analysis of over 400K target-disease pairs identified putative causal effect of levels of six proteins on COVID-19 severity, where four of them, ENTPD5, OAS1, FCRL3, ICAM5, showed European-only effects, one of them, SERPINA1, showed African-only effect, and one of them showed bi-ancestry effect. The MR-PheWAS of these prioritised targets showed little evidence of effect on other complex traits, which implies that these targets are unlikely to have a major adverse effect on complex human diseases. To enable the evidence to be widely accessible for COVID-19 drug target research, we released all our MR results via the EpiGraphDB platform (http://epigraphdb.org/covid-19/ctda/).

The *ENTPD5* gene encodes the NTPase Ectonucleoside Triphosphate Diphosphohydrolase 5, an enzyme mediating extracellular catabolism. A specific role of ENTPD5 is to promote host protein N-glycosylation for proper protein folding (information from Open Targets). SARS-CoV-2 spike protein is extensively N-glycosylated and blocking viral N-glycan biosynthesis was shown to inhibit viral entry^27^. Our results suggested that both gene and protein expression levels of *ENTPD5* showed an effect on COVID-19 severity, which provides evidence to support the examination of the direct role of ENTPD5 inhibition on SARS-CoV-2 viral entry and, subsequently, in the prevention of severe COVID-19 in future studies.

The *FCRL3* gene is an immunoglobulin receptor which is an antigen that was expressed in B cells. Previous studies suggested that patients with autoantibody against B cells had lower frequencies of anti-SARS-CoV-2-RBD IgM^28^. Our study suggested the putative causal role of protein level of FCRL3 on COVID-19 severity in the general European population, which further enhanced the link between the two.

The *SERPINA1* gene is a serine protease inhibitor that also inhibit TMPRSS2. Some clinical trials Several clinical trials of SERPINA1 inhibitor have been initiated for COVID-19 treatment. In this study, we predict the effect of these ongoing trials using genetic evidence, and further suggested that the effect of SERPINA1 inhibition may show stronger effect in African ancestry. However, we also noticed that several studies have suggested that urgent action is needed for this target, as SERPINA1 inhibition is related to alpha1-antitrypsin deficiency, which could be a risk of covid-19^29,30^.

Two genes, ICAM1 and ICAM5 belongs to the intercellular adhesion molecule family, showed effects on COVID-19 severity in our study. Proteins encoded by these two genes bind to the leukocyte adhesion LFA-1 protein, which is related to immune response. Recent study suggested that ICAM5/TYK2 gene as associated with severe COVID19^31^. Another study further suggested ICAM1 as a potential prognostic indicator for COVD-19 infection^32^. Correctively, more investigation of the role of ICAM family on COVID-19 is need.

Some limitations of our analysis are worth noting. Whilst initiatives are underway to collect genetic information for COVID-19 patients (e.g. the COVID-19 host genetics initiative, https://covid-19genehostinitiative.net/), the recent initial GWASs of COVID-19 found few genetic association signals, which highlights the importance of sample size and statistical power, especially in African ancestry. Second, recent MR studies of COVID-19 identified a few gene targets associated with COVID-19^12,13,14,15,16^, but these studies downplayed consideration of potential biases in the data. For example, if we use a mixture of population samples as controls (which include unexposed individuals and asymptomatic and untested cases), we will not be able to distinguish the causal effects of being exposed to SARS-CoV2, infected by the virus, or progressing to severe COVID-19 after infection^18^. A GWAS using an unbiased population sample screened to detect infection of COVID-19 (e.g. from antibody tests) will be helpful to disentangle this issue, but such a GWAS does not yet exist. Our study excluded the five available COVID-19 GWAS datasets (with populations or self-reported data as controls) to minimise the influence of mixed controls. We only used hospitalised COVID-19 vs. non-hospitalised COVID-19 as the outcome for MR, which had the most reliable definition of cases and controls. However, given the GWAS of hospitalized COVID-19 vs. not hospitalized COVID-19 was still selected on the basis of COVID-19 status, collider bias could still be an issue (see **Figure S3**). To estimate the influence of potential collider bias, we conducted MR-PheWAS of the prioritised targets. The prioritised targets such as ENTPD5 did not show much evidence of a causal effect on major diseases (e.g. asthma), which suggested that gene/protein level change of these targets are not obviously acting through these disease on selection (e.g. COVID-19 testing). However, the targets prioritised by our study should still be carefully reviewed in future trials before clinical application. Third, the drug targets evaluated in this study were proxied using a limited number of instruments, which means the putative causal effects rely on one or two genetic instruments. Thus, these associations support causality but do not prove it, as horizontal pleiotropy remains an alternative possibility. Fourth, even though our results suggest some biological links between the target and diseases, these only provide evidence for the very first step of the drug development process. Fifth, whilst these are plausible targets for COVID-19 severity, we cannot predict whether successful intervention would impact on risk of infection or other disease characteristics relevant to public health (e.g. viral shedding).

In conclusion, this study prioritised six proteins as potential drug targets for COVID-19 severity using an integrated genetic approach. Five proteins showed ancestry-specific effects, which evidences the value of genetic approaches in predicting drug response in different populations. This provides the very first step towards evaluating intervention targets that are worth following-up for all types of coronaviruses.

## Data Availability

The data needed for the analysis were available via MR-Base platform.
The results is available via the EpiGraphDB platform http://epigraphdb.org/covid-19/ctda/

## Supplementary Materials

**TableS1. The available pQTL instruments of 1,002 plasma proteins in European and African ancestries**.

**TableS2. The available eQTL instruments of 16,059 blood transcripts in European ancestry**.

**TableS3. Drugs in trials for COVID-19 treatment and their target genes**.

**TableS4. The available genetic information for SARS-CoV-2 target genes from pQTL and tissue specific eQTL resources**.

**TableS5. The genetic instruments and association information of the SARS-CoV-2 target genes in European ancestry**.

**TableS6. The outcome information of the genome-wide association studies of the 6 COVID-19 phenotypes**.

**TableS7. The list of 622 human phenotypes been used as outcomes for the phenome-wide mendelian randomization analysis**.

**TableS8. The plasma protein-COVID-19 severity associations with suggestive Mendelian randomization evidence (P<0.05) in European and African ancestries**.

Note: The target-COVID-19 severity associations with Mendelian randomization p value < 0.05 were included in this table.

**TableS9. The blood transcription-COVID-19 severity associations with suggestive Mendelian randomization evidence (P<0.05) in European ancestry**.

Note: The target-COVID-19 severity associations with Mendelian randomization p value < 10^−2^ were included in this table.

**TableS10. The target-COVID-19 severity associations with suggestive Mendelian randomization evidence (P<0.05)**.

**TableS11. The colocalization results for proteins, expression levels of ENTPD5 and COVID-19 severity**.

Note: The target-disease associations passed Bonferroni corrected threshold (6.0×10^−5^) were tested further by colocalization analysis.

**TableS12. The phenome-wide Mendelian randomization results for the 12 prioritised drug targets**.

**Figure S1. Mendelian randomization and colocalization models**.

Note: Model 1 – Causality: A genetic variant affects disease risk by changing protein levels; Model 2: Reverse causality Genetic variants affect disease risk through pathways other than via the protein of interest. The disease has a downstream effect on protein levels; Model 3 – Horizontal pleiotropy: a genetic variant influences both protein levels and disease risk by two independent biological pathways; Model 4 – confounding by LD: a genetic variant (variant 1) that influences protein levels is correlated with a second variant (variant 2) that influences disease risk. Colocalization analysis can distinguish Model 4 from Model 1 or Model 3.

**Figure S2. Manhattan plot demonstrating phenome-wide Mendelian randomization associations of protein expression level of ENTPD5 on human phenotypes**. Different colors refer to different disease areas. For the 622 tested traits, none of them passed the Bonferroni-corrected threshold is 8.04×10^−5^. Therefore, ENTPD5 are less likely to be influenced by horizontal pleiotropy.

**Figure S3. Potential influence of collider/selection bias on the MR association of a target on COVID-19 severity**.

Note: A collider could induce a spurious association between a protein and COVID-19, which affect the likelihood of an individual being sampled. As an example, COVID-19 testing could be a shared consequence (collider) of a disease (e.g. asthma) and COVID-19 severity, since people with respiratory illness are more likely to seek COVID-19 testing, as are people with more severe symptoms. This could induce a spurious association between asthma-associated protein levels and severity of COVID-19.

## Acknowledgments

This research was conducted using the UK Biobank resource under application number 15825. The UK Biobank received ethnical approval from the research ethnic committee (REC reference for UK biobank 11/NW/0382) and participants provided written informed consent.

## Author contributions

JZ and YMZ selected the drug targets; JZ performed the Mendelian randomization analysis; JZ and DB performed the colocalization analysis; JZ conducted the triangulation between MR and drug trials; JZ and YMZ conducted the drug target prioritisation; YL developed the database and web browser; JZ and YMZ wrote the manuscript; DB, YL, LW, XZL, HZ and TRG reviewed the paper and provided key comments; JZ, YMZ and TRG conceived and designed the study and oversaw all analyses.

## Statistics and Reproducibility

We have made all MR results openly available to browse or download at the COVID-19 Target-Disease Atlas (CTDA) browser within the EpiGraphDB platform (http://epigraphdb.org/covid-19/ctda/). This includes 14,873 unique target-COVID-19 severity associations evidence for the omics MR as well as 372,830 unique target-disease associations evidence for 353 targets on 622 diseases/phenotypes in 11 SARS-CoV-2 related tissues. Users are able to query the study results by the targeted gene/protein name and QTL SNPs via the online platform, and the results are presented in searchable tables as well as volcano plots. In addition, users can programmatically access the results using the /covid-19/ctda endpoints in the application programming interface (API) of EpiGraphDB via http://api.epigraphdb.org/.

To support reproducibility of our pipeline, we have made an Github repository to share the script of running Mendelian randomization and follow-up analyses. https://github.com/MRCIEU/epigraphdb-ctda

