## Supplemental Figure for "Multi-ancestry omic Mendelian randomization revealing putative drug targets of COVID-19 severity"

### Supplementary Figures

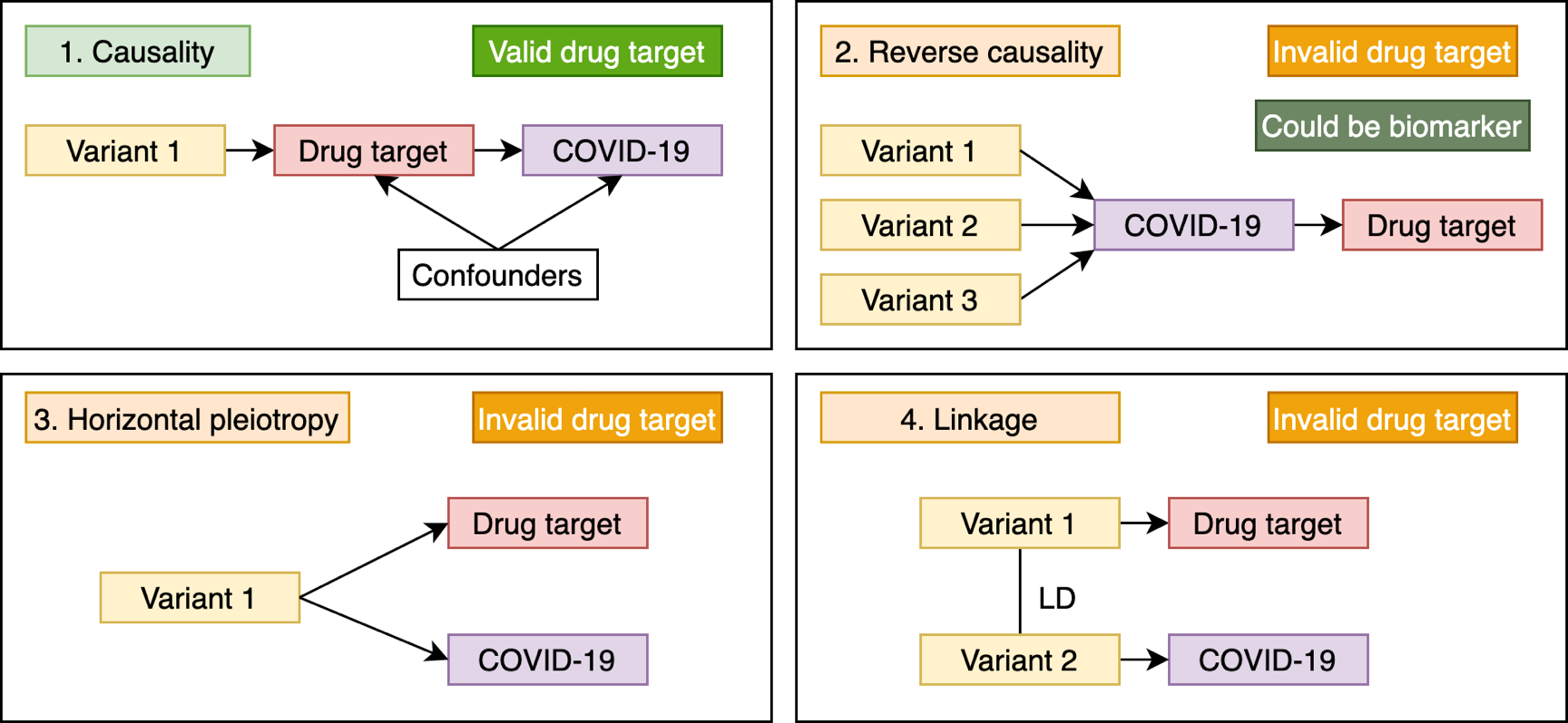

**Supplementary Figure 1. Mendelian randomization and colocalization models.** Model 1 – Causality: A genetic variant affects disease risk by changing protein levels; Model 2: Reverse causality Genetic variants affect disease risk through pathways other than via the protein of interest. The disease has a downstream effect on protein levels; Model 3 – Horizontal pleiotropy: a genetic variant influences both protein levels and disease risk by two independent biological pathways; Model 4 – confounding by LD: a genetic variant (variant 1) that influences protein levels is correlated with a second variant (variant 2) that influences disease risk. Colocalization analysis can distinguish Model 4 from Model 1 or Model 3.

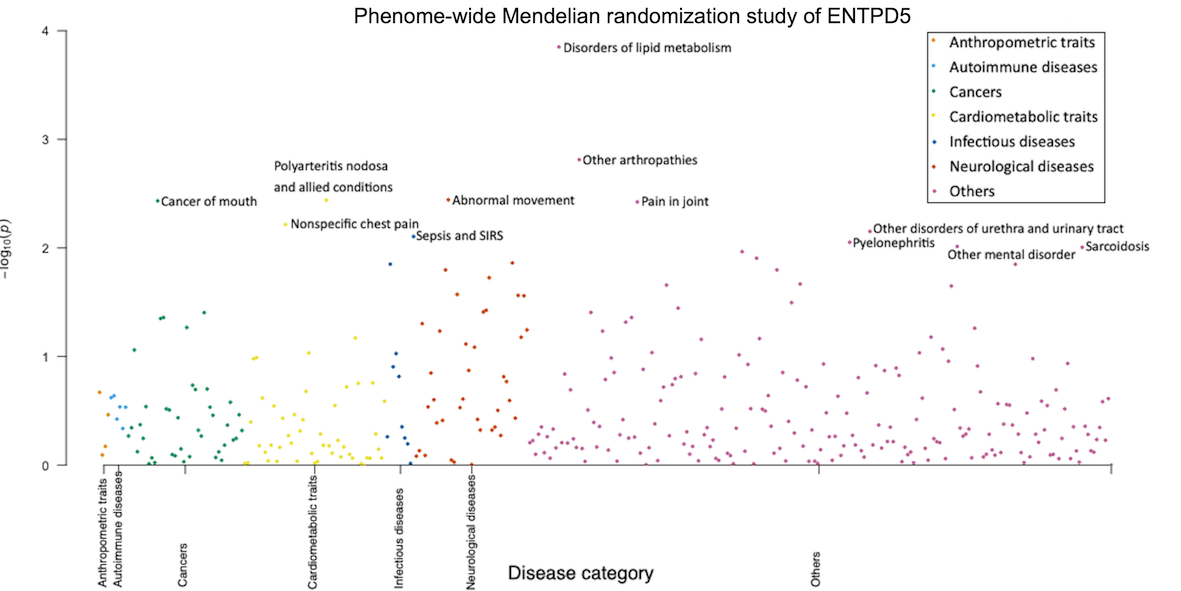

**Supplementary Figure 2. Manhattan plot demonstrating phenome-wide Mendelian randomization associations of protein expression level of ENTPD5 on human phenotypes.** Different colors refer to different disease areas. For the 622 tested traits, none of them passed the Bonferroni-corrected threshold is 8.04×10^-5^. Therefore, ENTPD5 are less likely to be influenced by horizontal pleiotropy.

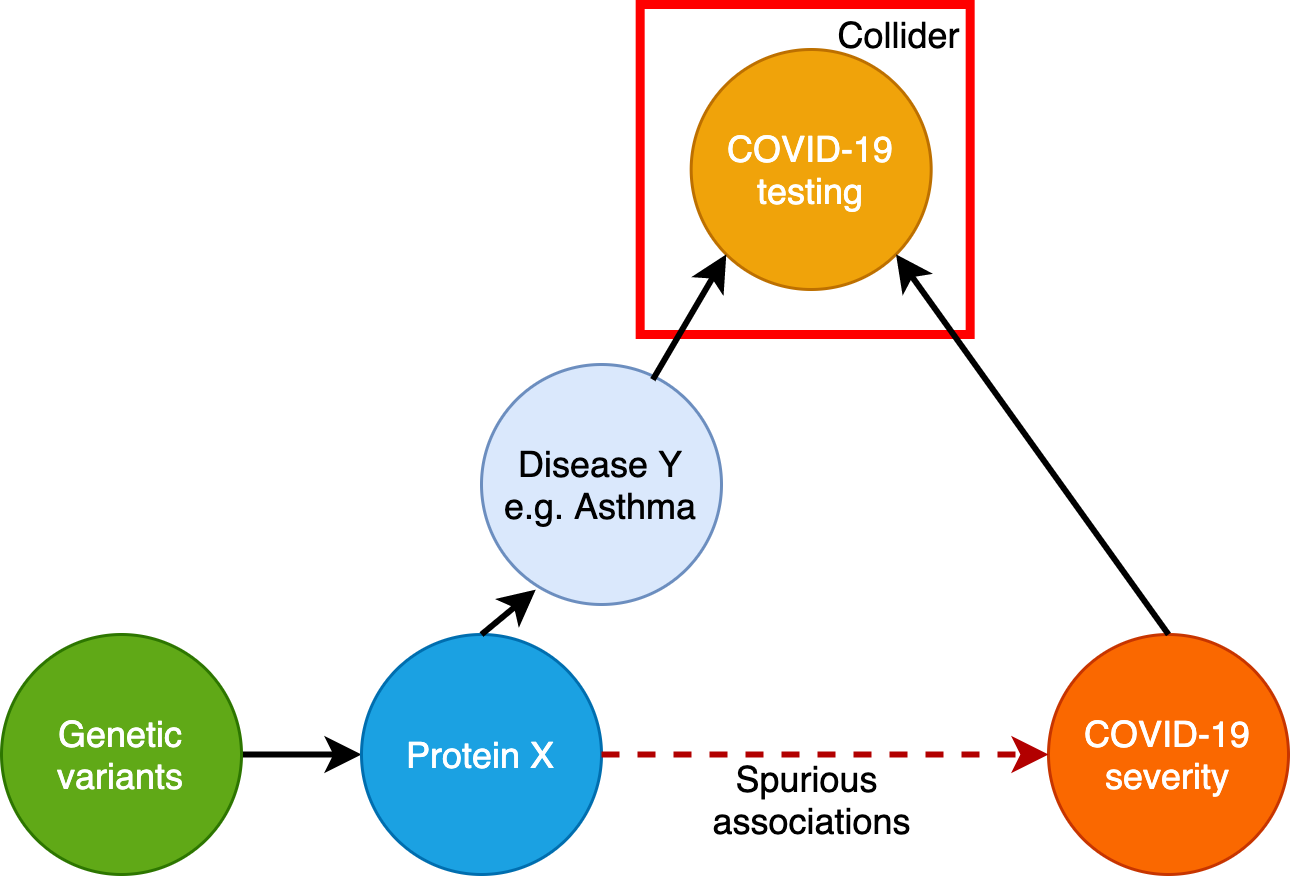

**Supplementary Figure 3. Potential influence of collider/selection bias on the MR association of a target on COVID-19 severity.** A collider could induce a spurious association between a protein and COVID-19, which affect the likelihood of an individual being sampled. As an example, COVID-19 testing could be a shared consequence (collider) of a disease (e.g. asthma) and COVID-19 severity, since people with respiratory illness are more likely to seek COVID-19 testing, as are people with more severe symptoms. This could induce a spurious association between asthma-associated protein levels and severity of COVID-19.

### Note S1. Instrument selection

First, genetic association information of gene and protein expression levels were looked up from two resources: i) protein expression levels in plasma from five studies ^12345^ implemented in Zheng et al. ^6^ ; ii) gene expression levels in whole blood from eQTLGen consortium ^7^. The genetic variants, genes and proteins were further mapped to genome build GRCh37.p13 coordinates. We used the following criteria to select genetic instruments:

1. We selected SNPs that were associated with any protein or gene expression (using a P-value threshold ≤5x10^-8^) in at least one of the four GWASs, including both cis and trans instruments.
2. We then conducted LD clumping for the instruments using the 1000 Genome European samples as reference panel, which was implemented in the TwoSampleMR R package (14) to identify independent instruments for each protein/gene. We used r^2^ < 0.001 as the threshold to exclude correlated instruments in the cis (or trans) gene region.

For multi-ancestry phenome-wide MR, pQTLs associated with plasma proteins from the Atherosclerosis Risk in Communities Study (ARIC) were used as instruments. All conditionally independent pQTLs at a false discovery rate (FDR) < 0.05 and LD r^2^<0.6 were kept for selection, with 6,614 pQTLs of 1,310 proteins in 7,212 Europeans and 3,900 pQTLs of 1,311 proteins in 1,871 Africans for MR analysis. F-statistics > 10 was used to ensure the instrument strength. Also, Steiger filtering was used to test the directionality of the pQTL-disease associations for all candidate instruments. Any pQTLs with Steiger filter flag as FALSE were removed. More details of the instrument selection process were presented in previous trans-ethnic paper^8^.

After instrument selection, 1,674 conditioning distinguished pQTL signals of 1,002 proteins (**Table S1**) and 39,630 conditioning distinguished eQTLs signals for 16,059 transcripts (**Table S2**) were kept as instruments for the genetic analyses of this study. To identify cis and trans instruments, we split instruments into two groups: 1) 29,610 cis-acting instruments within a 500Kb window from each side of the leading QTL of the protein/transcript; 2) 10,020 instruments outside the 500Kb window of the leading QTL were designated as trans instruments. Whilst trans instruments may be more prone to pleiotropy, their inclusion could increase statistical power as well as the scale of the study. Therefore, for the proteins and genes with cis instruments or trans instruments, we conducted MR analyses using both sets of instruments (**Table S1** and **S2**).

Second, we selected additional drug targets that were prioritised by evidence from the following three resources: i) a list of 11 drugs under trials for COVID-19 treatment were extracted from *ClinicalTrials.gov* (**Table S3**). These drugs were mapped to their target genes using Drug-Gene-Interaction (DGI) database (<http://dgidb.org/>) ^9^ and CHEMBL database ^10^; ii) human proteins interacting with SARS-CoV-2 proteins from Gordon et al. ^11^; iii) genes associated with SARS-CoV from Gralinski et al. ^12^. After de-duplicating these, 380 unique drug targets were selected for our study (**Table S4**). Next, gene and protein expression levels of these targets were looked up from four resources: protein expression levels in plasma from four studies ^12345^ implemented in Zheng et al. ^6^, gene expression levels in whole blood from eQTLGen consortium ^7^, tissue specific gene expression levels in 7 tissues from the GTEx consortium ^13^ and gene expression levels in two kidney tissues from Gilles et al. ^14^. After the mapping step, 353 drug targets with genetic variants robustly associated with the transcripts and/or proteins were included as the start point of the instrument selection (**Table S5**).

For any of the 353 targets, we mapped the drug targets with related drug names using four platforms: OpenTargets ^15^, ChEMBL ^10^, DrugBank ^16^ and DGI platforms ^9^. After this analysis, we mapped one target, PLOD2, to the drugs which target it (**Table S9**). We further mapped the targets to the previously reported “druggable genome” ^17^. This study stratified the potential drug targets from across the genome into three tiers. Tier 1 (1427 genes) included efficacy targets of approved small molecules and biotherapeutic drugs, as well as targets modulated by clinical-phase drug candidates; tier 2 was composed of 682 genes encoding proteins closely related to drug targets, or with associated drug-like compounds; and tier 3 contained 2370 genes encoding secreted or extracellular proteins, distantly related proteins to approved drug targets. For the 11 additional prioritised COVID-19 drug targets, one target was mapped to tier 1, 2 targets to tier 3 (**Table S9**).
